# Estimating the Impact of Control Measures to Prevent Outbreaks of COVID-19 Associated with Air Travel into a COVID-19-free country: A Simulation Modelling Study

**DOI:** 10.1101/2020.06.10.20127977

**Authors:** Nick Wilson, Michael G Baker, Martin Eichner

**Affiliations:** BODE^3^ Programme, University of Otago Wellington, New Zealand; HEIRU, University of Otago Wellington, New Zealand; Epimos GmbH, Germany; University of Tübingen, Germany

## Abstract

**Aims:** We aimed to estimate the risk of COVID-19 outbreaks associated with air travel from a country with a very low prevalence of COVID-19 infection (Australia) to a COVID-19-free country (New Zealand; [NZ]), along with the likely impact of various control measures for passengers and cabin crew.

**Methods:** A stochastic version of the SEIR model CovidSIM v1.1, designed specifically for COVID-19 was utilized. It was populated with data for both countries and parameters for SARS-CoV-2 transmission and control measures. We assumed one Australia to NZ flight per day.

**Results:** When no interventions were in place, an outbreak of COVID-19 in NZ was estimated to occur after an average time of 1.7 years (95% uncertainty interval [UI]: 0.04-6.09). However, the combined use of exit and entry screening (symptom questionnaire and thermal camera), masks on aircraft and two PCR tests (on days 3 and 12 in NZ), combined with self-reporting of symptoms and contact tracing and mask use until the second PCR test, reduced this risk to one outbreak every 29.8 years (0.8 to 110). If no PCR testing was performed, but mask use was used by passengers up to day 15 in NZ, the risk was one outbreak every 14.1 years. However, 14 days quarantine (NZ practice in May 2020), was the most effective strategy at one outbreak every 34.1 years (0.86 to 126); albeit combined with exit screening and mask use on flights.

**Conclusions:** Policy-makers can require multi-layered interventions to markedly reduce the risk of importing the pandemic virus into a COVID-19-free nation via air travel. There is potential to replace 14-day quarantine with PCR testing or interventions involving mask use by passengers in NZ. However, all approaches require continuous careful management and evaluation.

## Introduction

The COVID-19 pandemic has had major international health impacts during 2020, with 6.3 million cases and 380,000 deaths globally by 3 June (*1*). In many countries, border controls have been used to limit pandemic spread and this (combined with fear of the pandemic) have markedly reduced international travel. This reduction in travel has contributed to adverse economic and social impacts for countries by reducing business interactions, tourism and movements of international students.

New Zealand is one of the few countries that has eliminated transmission of the SARS-CoV-2 pandemic virus within its borders in line with the goal it adopted to achieve this (*2*). Some Australian states may also be approaching elimination status, but for Australia as a whole, elimination might not be achievable and the country might persist with a suppression strategy until a vaccine is widely available. Nevertheless, quarantine-free travel between the two countries is a goal envisaged by the Prime Ministers of Australia and New Zealand in terms of a trans-Tasman “bubble” (*3*). Such an approach has been discussed by the leaders of Austria, Greece, Israel, Norway, Denmark, the Czech Republic, Singapore, Australia, and New Zealand. These leaders “agreed that as each begins to ease restrictions they could capitalise on low infection rates by creating tourism safe zones” (*4*).

Nevertheless, such travel arrangements may be even more likely between New Zealand and those Pacific Island nations that have either successfully eliminated COVID-19 (e.g., as Fiji has declared) or which have been able to keep it out entirely due to strict border controls (e.g., Samoa, Tonga and Vanuatu).

Another development has been that many major airlines are also bringing in procedures to improve safety on flights to reduce the risk of SARS-CoV-2 transmission. These include physical distancing procedures and requirements for passengers and cabin crew to wear masks (*5*).

Given this background, we aimed to estimate the risk of COVID-19 outbreaks associated with increased air travel from Australia to New Zealand, along with the likely impact of various control measures that could be used to minimize the risk of such outbreaks.

## Methods

### Model design and parameters for SARS-CoV-2 and COVID-19

We used a stochastic SEIR type model with key compartments for: susceptible [S], exposed [E], infected [I], and recovered/removed [R]. The model is a stochastic version of CovidSIM which was developed specifically for COVID-19 (http://covidsim.eu; version 1.1). Work has been published from previous versions of this model (*6*) (*7*), and two preprints detail the equations and their stochastic treatment (*8, 9*). The model was built in Pascal and 100 million simulations were run for each set of parameter values. Such a large number of simulations was necessary due to the high probability of zero infected individuals on a flight given the low prevalence of infection in Australia (see below).

The parameters were based on available publications and best estimates used in the published modelling work on COVID-19 (as known to us on 27 May 2020). A key one was that 65% of infected COVID-19 cases develop clearly detectable symptoms (Table 1). Another was the effective reproduction number (Re) in COVID-19-free New Zealand, which was assumed to be 2.0 (Table 1).

**Table 1:** Input parameters used for modelling the potential spread of COVID-19 infections with the stochastic version of CovidSIM (v1.1) with New Zealand as a case study

| Parameter | Value/s used | Further details for parameter inputs into the modelling |
| --- | --- | --- |
| Latency period | 5 days | We used the best estimate from CDC in May 2020 of a mean of 6 days to symptoms (i.e., the latency period plus the prodromal period) (19). We used a standard deviation (SD) of 25% (1 day) (calculated using 16 stages; Erlang distribution). |
| Prodromal period | 1 day | There is, as yet, insufficient information on this prodromal period for COVID-19, so we used an assumed value for influenza (SD = 25%; 0.25 days, Erlang distribution). |
| Symptomatic period | 10 days (split into 2 periods of 5 days each) | The WHO-China Joint Mission report stated that “the median time from onset to clinical recovery for mild cases is approximately 2 weeks and is 3-6 weeks for patients with severe or critical disease” (20). But given that mild cases may have been missed in this particular assessment, we used a slightly shorter total time period of 10 days (SD = 25%; 2.5 days, Erlang distribution). |
| Infections that lead to sickness (symptomatic illness) | 65% | We used the best estimate from CDC in May 2020 of 65% symptomatic and 35% asymptomatic (19). This is more than the 20% proportion of asymptomatic cases as per a Chinese study (21), and as used in an Australian modelling study (22). This value is, however, lower than the 43% found in an Icelandic study (23). But it is also higher than that found in another Chinese study (at 6% asymptomatic) (24). A UK study of a cohort of health care workers reported that 27% of all infections were asymptomatic – but this group will be of different ages than the general population (25). |
| **Contagiousness** |  |  |
| Effective reproduction number (Re) in the NZ post-pandemic setting | 2.0 | We considered the best estimate from CDC in May 2020 of $R_0 = 2.5$ (19) but then reduced this to 2.0 to account for behavioral changes in the post-pandemic period (i.e., after the first wave in February to April 2020) in NZ. That is such changes as: some persisting voluntary physical distancing such as more working from home, higher levels of online shopping, enhanced hygiene behaviors, and some residual caution around attending large events. |
| Relative contagiousness in the prodromal period | 100% | We used the best estimate from CDC in May 2020 of infectiousness of asymptomatic individuals relative to symptomatic individuals of 100% (19). |
| Contagiousness after the prodromal period | 100% and 50% | In the first five days of symptoms, cases were considered to be fully contagious. In the second five-day period, this was assumed to be at 50%. The latter figure is still uncertain, but is broadly consistent with one study on changing viral load (26). |

### Prevalence of infection in Australia

To estimate the prevalence of SARS-CoV-2 infection in Australia, we assumed that there was the same number of undetected cases as there were detected cases. So we used 27 May 2020 data, when Australia reported 65 new detected cases for the preceding seven days (*10*). Then assuming a 16-day long period (latent period plus infectious period), this suggested a point prevalence of infected cases of 0.0006% ((65/7 × 16) / 25.46 million people). For the simulations, passengers were randomly sampled from the Australian population. The infection risk for the cabin crew was elevated due to in-flight transmission on serial flights that we modelled (detailed further below). In most of our scenarios, passengers and cabin crew members underwent entry screening before boarding (see Figure 1 and details below).

**Figure 1:**
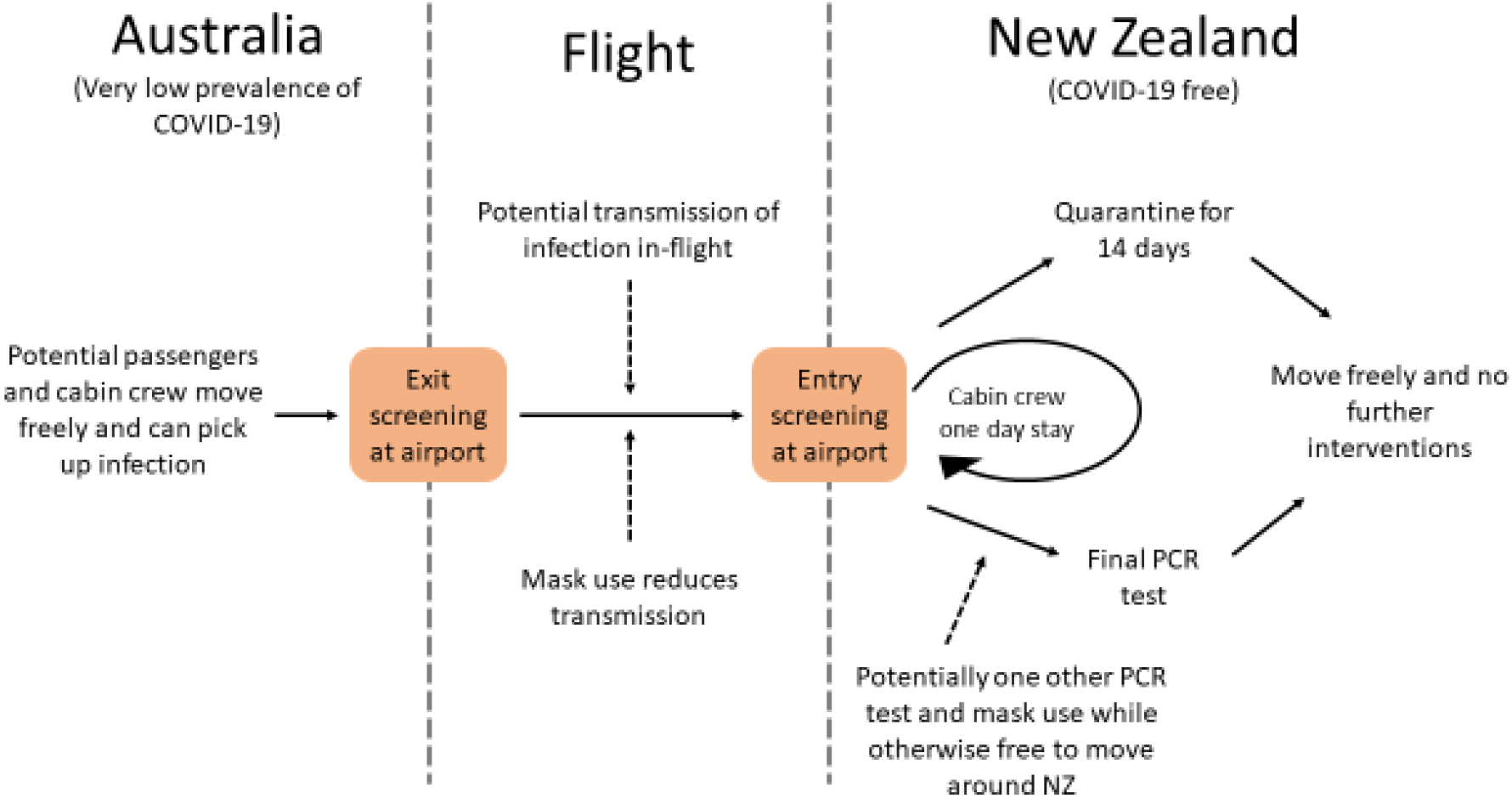
Flow diagram of the assumed movements of passengers and cabin crew in the model including interventions (simplified and not showing the precise details for how the cabin crew move back and forward between countries and the details around passengers seeking medical attention when symptomatic in New Zealand, isolation of identified cases and contact tracing)

### Selection of control measures

We identified plausible control measures from the published literature and also an online review of strategies identified by an IATA Medical Advisory Group (*11*). These controls are shown in Figure 1 and Table 2.

**Table 2:** Control measures used and their estimated efficacy in preventing SARS-CoV-2 transmission

| Control measure | Key value | Comment |
| --- | --- | --- |
| Airport **exit screening** for COVID-19 for both passengers and cabin crew leaving Australia (symptom questionnaire and thermal camera) | 50% of symptomatic cases are identified | Many infected passengers will be asymptomatic (including presymptomatic) at the time of exit screening. E.g., a modelling study of COVID-19 estimated that only 44% (95%CI: 33 to 56) of infected cases would be detected by exit screening (27). But this study only considered thermal camera scanning (with a sensitivity of 86% (28)) and did not consider a symptom questionnaire as well. |
| Required **mask use** on flights by passengers and cabin crew (except when drinking and eating) | 90% efficacy for both groups | We considered the results of a systematic review and meta-analysis that reported that mask use could result in a large reduction in risk of infection (n=2647; adjusted odds ratio = 0.15, 95%CI: 0.07 to 0.34) (29). But we used a slightly higher estimate to better approximate supervised and enforced mask use on the aircraft flight.<br>Of note is that many major airlines are now requiring mask use by passengers (5), and the use of masks on planes by the general public is encouraged by the World Health Organization (30). Also of note is the experimental evidence for mask efficacy. E.g., one experimental study using Avian Influenza virus to simulate the pandemic coronavirus, showed that: "N95 masks, medical masks, and homemade masks made of four layer kitchen paper and one layer cloth could block 99.98%, 97.14%, and 95.15% of the virus in aerosols" respectively (31). Another experiment has shown how a damp cloth over a speakers mouth reduces emitted droplets detected by laser light scattering by over 99% (32). More specifically another study reported that a non-fitted surgical mask was 100% effective in blocking detection of seasonal coronavirus (albeit slightly less effective for when measuring viral load in the samples) (33). |
| **Entry screening** on arrival in NZ (both passengers and cabin crew) | 50% of symptomatic cases identified | As per exit screening above (i.e., symptom questionnaire and thermal camera). Unless stated otherwise, we always applied entry screening for crew members; for passengers, we only used entry screening if they were not immediately to have a PCR test on arrival or are quarantined anyway. |
| **Quarantine** in NZ for passengers only (current practice as per May 2020 in NZ) | 14 days | We ran the simulations for the 14-day quarantine period to determine how many passengers were still infectious when quarantine ended. Of note is one estimate (34) that around 1% of people will still develop symptoms after 14 days (and will also be infectious at this time), but we do not use this estimate in our simulations, because the correct fraction depends on the infection stage of passengers at the beginning of their quarantine. |
| **Testing instead of 14 days quarantine:** PCR test for SARS-CoV-2 at various times | The time course of sensitivity values from Kucirka et al. was used | We used the results of a study (35) which fitted a Bayesian hierarchical logistic regression model for test sensitivity. This meant for example, at day 4 after infection, 67% of test results were false negatives (95%CI: 27% to 94%). This decreased to 20% (95%CI: 12% to 30%) on day 8 and then increased after this e.g., up to 66% (95%CI: 54% to 77%) on day 21. For cases who already recovered before their PCR test, we use the final value reported by Kucirka et al. (i.e., 34% sensitivity). In the days after arrival and before the next PCR test, we assume that people act normally and so can potentially spread infection to the NZ public (albeit with mask use when with other people). For more details, see Appendix Two.<br>In the absence of relevant data, we had to assume test result independence i.e., a false negative for a test was not correlated with a false negative for a later test. If both results were negative, we assumed no further follow-up.<br>We considered a wide range of different timing options for PCR tests after arrival in NZ. |
| Contact tracing if (i) a scheduled PCR test is positive or (ii) if people develop symptoms and seek medical attention (see below) | 75% of infected contacts are traced and isolated | No detailed data on the efficacy of contact tracing has been published for NZ. Nevertheless, the success of contact tracing to identify the source of clusters as detailed on the NZ Ministry of Health website is 62% (10/16 clusters, 30 May data (36)). However, we assume here a slightly higher success rate given on-going learnings by the NZ health system. |
| Mandatory mask use by incoming | 90% transmission | See above for details on the likely efficacy of mask use in reducing contagiousness (i.e., from an infected person spreading infection to |
| passengers up to the time of their final PCR test | reduction (as per above) but also no mask use in a scenario analysis | others). We also used a higher efficacy than reported in the systematic review (29) (detailed above), on the basis that this group of passengers would be provided information on the flight and on arrival in NZ on the critical importance of mask use and its mandatory nature, along with a free supply of masks on arrival). |
| The proportion of infected passengers who when they develop any symptoms seek medical attention (i.e., they are in the 65% who will ever develop symptoms) | 50% (self-reporting occurs on average 1 day after symptom onset) | We assumed that this proportion is somewhat higher than that for the general community (see below) on the basis that these passengers would be provided information on the flight and on arrival in NZ on the critical importance of seeking medical attention if they develop any symptoms. They would also be told that such medical attention would be provided free of charge. We assumed PCR confirmation of self-reported symptoms and if a positive test, then we assumed case isolation and potentially triggering contact tracing.<br>Of note is that routinely in NZ, 39.5% of people with “fever and cough” symptoms seek medical attention, as reported by the NZ Flutracking surveillance system (37). This is very similar to international estimates for people with influenza who seeking medical attention at 40% e.g., as used in other modelling (6). |
| Quarantine of traced contacts | 1 day after detection of index cases | Traced contacts are assumed to be effectively quarantined with no further spread of infection. |

### Air travel from Australia to New Zealand

We simulated one flight per day from Australia to New Zealand, carrying 300 passengers and 6 cabin crew members. A wide range of aircraft were used on this route in the pre-pandemic era with common ones being the Boeing 777-200 which takes 312 passengers and the Airbus A300-300 with 297 passengers. We used the minimum ratio of one cabin crew member to 50 seats (as required by some regulators), on the assumption that there might be new processes that reduce crew workloads (e.g., meals/drinks placed on seats in advance). This is a small proportion of the level of travel in the pre-pandemic time (i.e., 7.1% of the of 1,542,467 visitor arrivals from Australia to New Zealand in the year to January 2020 (*12*)).

### In-flight transmission risk

There are several publications that suggest transmission of SARS-CoV-2 on aircraft. One reported on 11 patients who “were diagnosed after having flown together in the same flight with no passenger that could later be identified as the source of infection” (*13*). Another reported a single case “most likely acquired during a flight” (*14*). But a flight with an index patient who had a dry cough when onboard did not appear to spread infection to any of the approximately 350 passengers (*15*). An IATA document (*11*) has reported a number of flights with passengers with SARS-CoV-2 infection who apparently infected no other passengers, but also one UK to Vietnam flight with “up to 14 people” infected. Also, based on an IATA survey of four airlines, it was reported that: “There was one possible secondary passenger case identified in the total, along with just two crew cases, thought to be the result of possible in-flight transmission” (*11*).

Also, of note is that there has been a COVID-19 outbreak in Zhejiang Province, China on a bus (*16*): passengers on the same bus as the index case had an infection risk ratio of 41.5 (95% CI: 2.6–669.5) compared with passengers on another bus. There has also been an aircraft outbreak of SARS (the coronavirus SARS-CoV-1) where one symptomatic person generated 16 laboratory-confirmed secondary cases (with 6 other likely cases) (*17*).

Given this background, it is obviously difficult to quantify the risk on SARS-CoV-2 transmission on board of a passenger aircraft. Therefore, for simulation purposes, we used quantified data from influenza transmission on aircraft. To do this we extracted data from a relevant systematic review and re-analyzed it (see Appendix One). This work indicated that an index case might typically generate an average of 6.0 secondary cases on a flight of at least three hours duration (assuming no mask use and assuming SARS-CoV-2 is similar to influenza in infectivity in such situations). We assumed that cabin crew had the same per person risk of being infected and the same transmission rate as any of the passengers (in the absence of any data on this from the systematic review). We also considered compulsory mask use on flights, as per major airlines in June 2020 (*5*).

### Arrival in New Zealand

Upon arrival in New Zealand, we assumed there is entry screening for both passengers and cabin crew. Passengers were either placed in supervised quarantined for 14 days (as per actual arrangements in May 2020) and then released to move freely, or, as an alternative to 14 days of quarantine, we considered various combinations of PCR testing in New Zealand. Indeed, the PCR test on arrival is already in use in some settings (i.e., Austria in May 2020) with a three hour waiting time until test results. Up until their last PCR test, we assumed that people could move freely around New Zealand but were required to wear a mask while in the presence of other people; we further assumed that half of the cases who develop symptoms during this period would report these symptoms within one day. Also, we assumed that if these people are tested positive, or if they reported symptoms themselves, contact tracing would identify 75% of their infected contacts in New Zealand who would be isolated after another delay of one day.

After entry screening, cabin crew arriving in New Zealand are not quarantined but are assumed to stay for one day in New Zealand before their next flight (albeit in an scenario analysis they stay in special facilities and do not mix with the public as per some existing processes for New Zealand (*18*)).

### Ongoing infection transmission in New Zealand

Secondary cases who were infected by crew members or passengers in New Zealand, yet who were not traced, and tertiary cases who are infected by traced secondary cases before they were isolated, were assumed to have the full length of their infectious period ahead of them. Some of them then can trigger an outbreak.

### Return flight to Australia (cabin crew)

To make the simulations as realistic as possible, cabin crew members were assumed to travel back to Australia after their one day layover in New Zealand, taking any infection they previously acquired back with them. They were then assumed to have another layover in Australia of one day (where uninfected cabin crew members could also pick up the infection from interacting with the public) and then they may or may not be detected (and removed) at the next boarding screening process. While cabin crew were subjected to exit and entry screening and the wearing of masks onboard, these were the only interventions we considered for them (albeit scenario analysis regarding not having a one day layover in either country).

### Control measures assumptions

The full details on the control measures we considered are detailed in Table 2.

## Results

For the base case of one flight per day from Australia and where no interventions were assumed to be in place, we estimated that New Zealand would on average experience an outbreak of COVID-19 after 1.7 years (Table 3). This was increased to 2.2 years after adding in exit screening upon leaving Australia; to 3.3 years by adding in mask use on flights; and to 3.5 years by adding in entry screening on arrival in New Zealand. When adding in PCR testing (accompanied by self-reporting of symptoms, wearing of masks, and contact tracing/case isolation up to the last test), the various scenarios indicated average times to an outbreak of 4.4 to 29.6 years (Table 3). The key driver of these results was not the number of tests but the timing of the last test, as this extended the benefits of mask use, symptom reporting and contact tracing. Additional results with the time to last PCR test extended up to day 15 in New Zealand (Table 4), indicate the particularly important benefit from mask use, then symptom self-reporting, and then contact tracing/isolation. The best result was an average outbreak time of 29.6 years (Table 3). Nevertheless, if PCR testing was completely abandoned and replaced with just mask use up to day 15 in New Zealand, there was still a reasonable benefit (i.e., 14.1 years for “mask use after arrival” in Table 4).

**Table 3:** Results of the simulations of the base case risk (no interventions) and for multi-layered packages of interventions to prevent COVID-19 outbreaks in New Zealand (NZ) (assuming the prevalence of infection in Australia (AU) is 0.0006% as per our best estimate for May 2020).

| Strategy | Exit screening in AU (detection of sick cases) | All wear masks on board * (prevention) | Entry screening in NZ (detection of sick cases) | Quarantine of passengers | PCR tests for passengers (day 1 is arrival day)* | Contact tracing between PCR tests | Passengers wear masks in NZ until last PCR test ** | Self-reporting of symptoms *** | Self-reporting triggers contact tracing | Expected duration until outbreak in NZ (years) | Uncertainty interval in years (95%) |
| --- | --- | --- | --- | --- | --- | --- | --- | --- | --- | --- | --- |
| No PCR tests, no quarantine | - | - | - | - | - | - | - | - | - | 1.7 | 0.04-6.09 |
|  | 50% | - | - | - | - | - | - | - | - | 2.2 | 0.06-8.11 |
|  | 50% | 90% | - | - | - | - | - | - | - | 3.3 | 0.08-12.1 |
|  | 50% | 90% | 50% | - | - | - | - | - | - | 3.5 | 0.09-12.9 |
| **PCR tests (in addition to various impacts from exit and entry screening and masks on flights etc)** |  |  |  |  |  |  |  |  |  |  |  |
| PCR tests (see Table 4 for more details) | 50% | 90% | - | - | day 1 | - | - | - | - | 4.4 | 0.11-16.1 |
|  | 50% | 90% | - | - | days 1 + 8 | 75% | - | - | - | 6.4 | 0.16-23.7 |
|  | 50% | 90% | - | - | days 1 + 8 | 75% | - | 50% | 75% | 7.5 | 0.19-27.7 |
|  | 50% | 90% | - | - | days 1 + 8 | 75% | 90% | - | - | 17.0 | 0.43-62.7 |
|  | 50% | 90% | - | - | days 1 + 8 | - | 90% | 50% | - | 19.9 | 0.50-73.2 |
|  | 50% | 90% | - | - | days 1 + 8 | 75% | 90% | 50% | 75% | 20.0 | 0.51-73.6 |
|  | 50% | 90% | 50% | - | days 3 + 12 | 75% | - | - | - | 5.4 | 0.14-19.8 |
|  | 50% | 90% | 50% | - | days 3 + 12 | 75% | - | 50% | 75% | 6.4 | 0.16-23.6 |
|  | 50% | 90% | 50% | - | days 3 + 12 | 75% | 90% | - | - | 23.5 | 0.60-86.8 |
|  | 50% | 90% | 50% | - | days 3 + 12 | - | 90% | 50% | - | 28.1 | 0.71-104 |
|  | 50% | 90% | 50% | - | days 3 + 12 | 75% | 90% | 50% | 75% | 29.6 | 0.75-109 |
| **Quarantine (in addition to impacts from exit and entry screening and masks on flights etc)** |  |  |  |  |  |  |  |  |  |  |  |
| Quarantine | 50% | 90% | 50% | 7 days | - | - | - | - | - | 5.8 | 0.15-21.5 |
|  | 50% | 90% | 50% | 14 days | - | - | - | - | - | 34.1 | 0.86-126 |

\* A range of days were considered, but the 3 + 12 day option is the one currently adopted for all travelers as per June 2020 in NZ (albeit combined with quarantine for all travelers to NZ).
\*\* Prevention of secondary infections due to wearing of masks by potentially infected individual passengers.
\*\*\* The given fraction of passengers who report having developed symptoms while staying in NZ to the health system; they are assumed to be isolated one day after symptom onset and contact tracing may occur after this; traced contacts are PCR tested and isolated after another delay of one day.

**Table 4:** Expected duration (in years) until a COVID-19 outbreak in New Zealand for differing duration of intervention and for different timing of PCR tests for arriving passengers while in New Zealand (assuming exit screening in Australia and mask use on flights in all scenarios)

| Scenario | Intervention until day (counting arrival day as day 1 in NZ) |  |  |  |  |  |  |  |  |  |  |  |  |  |
| --- | --- | --- | --- | --- | --- | --- | --- | --- | --- | --- | --- | --- | --- | --- |
|  | 2 | 3 | 4 | 5 | 6 | 7 | 8 | 9 | 10 | 11 | 12 | 13 | 14 | 15 |
| 2 PCR tests<sup>*</sup>, self-reporting of symptoms, contact tracing, but passengers do not wear masks in NZ | 6.2 | 6.6 | 7.2 | 7.8 | 8.2 | 8.0 | 7.5 | 7.1 | 6.7 | 6.3 | 6.0 | 5.9 | 5.8 | 5.7 |
| Passengers wear masks in NZ<sup>**</sup>, but no PCR tests, no self-reporting of symptoms, no contact tracing | 3.6 | 3.8 | 4.0 | 4.2 | 4.5 | 4.9 | 5.4 | 6.0 | 6.9 | 7.9 | 9.2 | 10.4 | 12.2 | 14.1 |
| Passengers wear masks in NZ and self-report symptoms<sup>**</sup>, but no PCR tests and no contact tracing | 3.7 | 3.9 | 4.1 | 4.5 | 4.9 | 5.5 | 6.3 | 7.3 | 8.5 | 9.9 | 11.5 | 13.1 | 15.0 | 17.3 |
| Passengers wear masks in NZ, self-report symptoms with contact tracing<sup>**</sup>, but no PCR tests | 3.7 | 3.9 | 4.1 | 4.4 | 4.9 | 5.6 | 6.5 | 7.5 | 8.7 | 10.1 | 11.6 | 13.4 | 15.7 | 17.6 |
| Only 1 PCR test<sup>***</sup>, passengers wear masks in NZ and self-report symptoms, but no contact-tracing | 7.7 | 8.3 | 9.5 | 11.2 | 13.3 | 15.2 | 16.2 | 18.3 | 19.4 | 19.7 | 20.3 | 20.7 | 21.6 | 22.3 |
| 2 PCR tests<sup>1</sup>, passengers wear masks in NZ, but no self-reporting and no contact tracing | 7.0 | 7.7 | 9.0 | 11.0 | 13.5 | 16.0 | 17.2 | 18.6 | 19.5 | 19.8 | 20.4 | 20.9 | 21.6 | 22.3 |
| 2 PCR tests<sup>1</sup> with contact tracing, passengers wear masks in NZ but no self-reporting of symptoms | 6.9 | 7.8 | 9.0 | 11.0 | 13.6 | 16.1 | 17.3 | 18.8 | 19.7 | 20.1 | 20.6 | 21.3 | 22.2 | 23.4 |
| 2 PCR tests<sup>1</sup>, passengers wear masks in NZ and self-report symptoms, but no contact tracing | 8.1 | 8.9 | 10.4 | 12.5 | 15.2 | 17.8 | 19.8 | 22.6 | 23.8 | 24.5 | 25.4 | 25.8 | 26.4 | 27.7 |
| 2 PCR tests<sup>1</sup>, passengers wear masks in NZ and self-report symptoms; both, PCR tests and self-reporting trigger contact tracing | 8.2 | 9.0 | 10.4 | 12.4 | 15.5 | 18.5 | 20.4 | 23.3 | 24.9 | 25.6 | 26.3 | 26.9 | 28.0 | 29.5 |

Each result was obtained by running 100 million stochastic simulations.
\* First PCR is performed instead of entry screening immediately after arrival, second PCR is performed on the last intervention day; all interventions stop thereafter.
\*\* No PCR tests are performed; entry screening after arrival; all interventions stop after the last intervention day.
\*\*\* Entry screening after arrival; first PCR test occurs on the last intervention day; all interventions stop thereafter.

If 14-days of quarantine was used instead of PCR testing (as per actual practice in May 2020 in New Zealand), this would be by far the most effective intervention as it increased the average time to an outbreak to over 34.5 years for all the scenarios considered. Reducing the quarantine from 14 to 7 days resulted in a much more pessimistic result (average waiting time for an outbreak: 5.8 years).

Of note is that all these values are average durations until an outbreak occurs, yet each single flight bears an, albeit small, risk of triggering such an event. Outbreaks occur at a random time point (exponentially distributed) with the expected value given in Table 3. As the variance of the exponential distributions is extremely large, the 95% uncertainty intervals for when such an outbreak occurs are correspondingly very large. That is, in the most pessimistic scenario (no intervention scenario) with an expected duration of 1.7 years, 95% of outbreaks are predicted to occur between 0.04 years (15 days) and about 6.09 years; in the most optimistic scenario (with quarantine) with an expected duration of 34.1 years, 95% of outbreaks occur between 0.86 and 126 years.

Passengers as a group were the major driver of outbreak risk relative to cabin crew: they caused over 100 times as many outbreaks as cabin crew members (depending on the scenario). Passengers also caused more outbreaks on a per capita basis than cabin crew, given the 50:1 ratio of these groups per flight.

Table 5 indicates the extreme rarity of outbreaks in New Zealand if Australia’s infection prevalence was 10 times lower than in the base case (i.e., approximating if Australia is on the verge of elimination or has eliminated and then experiences a small undetected outbreak from a border control failure). But when scaling up from the base case to ten-fold higher infection prevalence in Australia or ten-fold increase in travel volumes from Australia to New Zealand, then the results scale up accordingly. For each of these scenarios individually, there would typically be an outbreak every 3.0 years for using PCR testing or 3.4 years for the quarantine intervention (Table 5).

**Table 5:** Results for major changes in the prevalence of infection in the source country (Australia) and flight volumes from Australia to New Zealand

| Parameters / scenarios | Expected duration until outbreak in NZ (years)<br>(95% uncertainty interval) |  |  |
| --- | --- | --- | --- |
|  | No interventions | PCR testing on days 3 + 12 | 14 days of quarantine |
| As per the base case. | 1.7 years<br>(0.04-6.09) | 29.6 years<br>(0.75-109) | 34.1 years<br>(0.86-126) |
| Australia's infection prevalence is 10 times lower than in the base case (i.e., 0.0006%). This could be when Australia is on the verge of elimination or has eliminated and then experiences an outbreak after a border control failure [Scenario A]. | 16.5 years<br>(0.42-60.9) | 296 years<br>(7.49-1090) | 341 years<br>(8.63-1260) |
| Australia's infection prevalence 10 times as high as in the base case (i.e., 0.006%). This could be due to poorer suppression or a winter upsurge [Scenario B]. | 0.17 years<br>(0.004-0.61)<br>[2.0 months] | 3.0 years<br>(0.07-10.9) | 3.4 years<br>(0.09-12.6) |
| Travel volume is 10 times as high as in the base case (i.e., 10 flights per day) with the infection prevalence as per the base case [Scenario C]. | 0.17 years<br>(0.004-0.61)<br>[2 months] | 3.0 years<br>(0.07-10.9) | 3.4 years<br>(0.09-12.6) |
| Combination of increased infection prevalence and increased travel volume as outlined above [Scenario B and Scenario C together]. | 0.02 years<br>(0.0004-0.06)<br>[6 days] | 0.3 years<br>(0.007-1.09)<br>[3.6 months] | 0.34 years<br>(0.01-1.26)<br>[4 months] |

100 million simulations for each set of parameter values. Values typically rounded to three meaningful digits.

## Discussion

### Main findings and interpretation

This analysis examined the risk of COVID-19 outbreaks in a COVID-19-free nation (New Zealand), if there was travel from a low-prevalence country (Australia) with no control procedures in place, and then if there were various multi-layered interventions in place. It suggested that without any controls there would be such an outbreak of COVID-19 in New Zealand after an average of 1.7 years (i.e., for just one flight per day from Australia and at a very low assumed prevalence of infection in Australia at just 149 undetected infected people in a population of 25 million). Fortunately, the multi-layered packages of interventions we have modeled can reduce this risk to much lower levels. In particular, quarantine for 14 days was by far the most effective single intervention. Nevertheless, the package of PCR testing (with mask use, symptom self-reporting and contact tracing) might be a reasonable alternative, with an expected outbreak occurring after an average of about 30 years.

Surprisingly, there was also a large benefit from just mask use alone for 15 days by arriving passengers. But specific complexities if quarantine is not used, include the issues of:

- Might PCR testing on arrival in New Zealand provide a false sense of reassurance to passengers if they test negative (and so subsequently impede their mask wearing adherence and self-reporting of symptoms to health workers)?
- Would the experience of PCR testing on arrival (if the taking of the nasopharyngeal sample is experienced as unpleasant), result in higher levels of defaulting for subsequent PCR testing?
- Might the requirement to turn up to a health provider for a PCR test on day 15, be an actual stimulus for improved mask adherence and symptom reporting (relative to intervention packages that did not involve any PCR testing)?
- Would adherence to mask use by passengers when in the New Zealand community be as high as we assume (given the minimal use of masks in the first pandemic wave in New Zealand and lack of Government mandates for mass masking at any of the four alert levels of COVID-19 response)? Also, might the use of fines and threat of deportation improve adherence with mask use amongst passengers once in New Zealand?
- Might digital tools for tracking and even active monitoring of early signs of illness have a role in minimizing risk amongst incoming passengers (especially in the first two to three weeks after arrival?

All these issues could benefit from further investigation, potentially involving state-of-the art study designs that remain possible under pandemic conditions (*38*).

Nevertheless, to select the best multi-layered package of interventions requires information on not only their estimated effectiveness, but also their estimated real-world cost-effectiveness, feasibility, acceptability and adherence with the traveling public, airlines and border control services. Obtaining such information would include studying the cost of dealing with false positives of PCR testing and the resources spent chasing up passengers who miss attending their scheduled PCR tests. There is also the need to identify the cost of the New Zealand public health system maintaining a state-of-the art contact tracing capacity (though this may be highly desirable for other reasons such as control of measles outbreaks). The relevance of these costs to policy-makers might also be impacted by who is paying. For example, all incoming passengers could be charged a COVID-19 levy and the whole system could be made user-pays and cost-neutral to taxpayers.

Ultimately, there is also a need for cost-benefit analyses which considers the benefits of increased travel to the New Zealand society and economy – along with the risks of outbreaks that need to be rapidly controlled (and might even get completely out of control). With such information policy-makers could then decide:

- On continuing with the 14-day quarantine or shifting to PCR testing with mask use interventions.
- On the extent to which they open up traveler volumes for different types of traveler from specified low-risk countries (e.g., including essential workers, students, business people, or tourists).
- On the extent to which they fund research on evaluating antibody tests to determine if some previously exposed passengers can be exempted some of the quarantine or PCR testing processes (though the quality of such antibody tests is still suboptimal and specificity might be a problem owing to other circulating coronaviruses).

### Study strengths and limitations

This is the first such modeling study (that we know of), to consider interventions to control SARS-CoV-2 spread by air travel between two countries. We were also able to consider a wide range of control interventions and to package these in multiple layers of defense and estimate uncertainty intervals. Nevertheless, there is quite high uncertainty around some of the parameters we used. For example, the prevalence of infection in Australia is highly variable by States/Territories, and this is also likely to vary over time. Real-world effectiveness of masks on aircraft is still uncertain, along with how well SARS-CoV-2 can spread on aircraft. For example, there is some evidence that this pandemic virus is particularly involved in super-spreading events with one estimate being that 80% of secondary transmissions may have been caused by a small fraction (e.g., ∼10%) of infectious individuals (*39*). Given all such issues and ongoing improvements in knowledge of the transmission dynamics of SARS-CoV-2, this type of modeling work should be regularly revised and be performed using different types of models.

## Conclusions

This analysis suggests that an outbreak of COVID-19 in New Zealand might occur after an average of 1.7 years without any interventions and for just one flight per day from Australia. This risk is greatly reduced by the currently used 14-day mandatory quarantine. Our analysis shows that there is potential to replace this quarantine period with multi-layered interventions using PCR testing or other controls, including mask use by passengers in New Zealand, that would also maintain a low risk of importing the pandemic virus. However, all approaches require continuous careful management and evaluation.

## Data Availability

All the relevant parameters used in the modelling are detailed within the article.

## Acknowledgements

Professor Wilson is supported by the New Zealand Health Research Council (16/443) and Ministry of Business Innovation and Employment (MBIE) funding of the BODE^3^ Programme (UOOX1406). Professor Michael Baker is supported by a New Zealand Health Research Council grant for research on COVID-19 (20/1066).

## Appendix One: Estimating the risk of SARS-CoV-2 in-flight transmission for one index case and with no mask use by cabin crew or passengers

### Methods

We extracted data from the single available systematic review of in-flight outbreaks of influenza (*40*), the majority (14/19) of which were related to the pandemic influenza of 2009. For one outbreak with missing flight duration data, we estimated this using Google Maps and one study was removed from the dataset as it appeared to be of the same flight as another included study (as noted by the review authors). The results of our analysis are shown in Table A1.

### Discussion

There is of course the limitation here that although pandemic influenza is a respiratory virus like SARS-CoV-2, these two agents might still have different transmission dynamics in the aircraft cabin setting. Another issue is that the in-flight influenza outbreaks in the systematic review did suffer from incomplete contact tracing (mean of 73% traced, median of 86% traced), and so the true burden of secondary cases is probably somewhat higher for these outbreaks. But countering this is the possibility that there might have been publication bias involved, given that in this review there were only 4/19 studies with zero secondary cases. This might suggest that large in-flight outbreaks of influenza were more likely to be studied (and published on) than those with no or little secondary spread. This may somewhat over-estimate the in-flight transmission of influenza, but it may be more appropriate for the transmission of SARS-CoV-2 which seems to have a higher reproduction number than influenza.

**Table A1:** Our analysis of extracted data from a published systematic review (*40*) of influenza outbreaks on aircraft flights

| Type of flight | Secondary cases per flight when just considering a single index case | Secondary cases per flight when considering both single and multiple index cases | Adjusted secondary cases per flight when scaling to the equivalent of a single index case in each outbreak |
| --- | --- | --- | --- |
| All flights (n = 19 flights*) | 8.0 | 8.3 | 5.2 |
| Flight length $\leq 3$ hours, $\leq 150$ passengers (n = 8 flights) | 7.6 | 7.8 | 5.3 |
| Flight length $\leq 3$ hours, $> 150$ passengers (n = 0 flights) | — | — | — |
| Flight length $> 3$ hours, $\leq 150$ passengers (n = 1 flights)* | Ignored: only 1 flight | Ignored: only 1 flight | Ignored: only 1 flight |
| Flight length $> 3$ hours, $> 150$ passengers (n = 10 flights)* | **6.0**<br>(this is the value we used for our Australia to NZ flight modeling for the no mask use scenario) | 7.6 | 3.7 |

\* Some longer flights included transfers and stopovers (which were treated in the analysis as one flight but we only considering flying time in the calculation of hours, except for one particular stopover where passengers remained on the plane).

## Appendix Two: Sensitivity of PCR tests during the course of infection

Kucirka *et al* (*35*) provide in their Figure 2 estimates on the time-course of the fraction of false-negative PCR results. We have reproduced their curve and translated it into a time-course that gives the test sensitivity for infected individuals, depending on the time since they were exposed to infection. As our simulations are not individual-based, but stochastic representations of a compartmental model, we cannot exactly know the time since exposure for an infected individual who is in a given state of infection (which is represented by one of the 16 latent stages, the 16 prodromal stages, the 16 early and the 16 late infections stages). In order to match these 4×16 infection stages, we ran one million stochastic simulations with one infected individual each, recorded the time when this individual entered or left any one of these stages and finally calculated the individual’s middle time point (since infection) for each of the 4×16 infection stages. In the next step, we recorded the sensitivity values that corresponded to the recorded 4×16 simulated time points of the individual, leading to one million simulation-matched values for each of the 4×16 infection stages. In a final step, we averaged over these one million values, obtaining 4×16 sensitivity values for the infection stages that we used in the simulations.

